# Physiotherapists Use Different Motivational Strategies Tailored to an Individual’s Condition: A Qualitative Study

**DOI:** 10.1101/2022.04.20.22274114

**Authors:** Kazuaki Oyake, Keita Sue, Motofumi Sumiya, Satoshi Tanaka

## Abstract

**Question:** How do physiotherapists use different motivational strategies for individuals in stroke rehabilitation?

**Design:** A qualitative study using in-depth semi-structured online interviews.

**Participants:** A criterion sample of 15 physiotherapists who have worked in rehabilitation for over 10 years and have an interest in an individual’s motivation.

**Intervention:** Not applicable.

**Outcome measures:** Physiotherapists’ perspectives and experiences regarding motivational strategies used depending on the individual’s condition.

**Results:** A total of nine themes emerged from the data upon thematic analysis and inductive coding. The participants used different strategies to encourage individuals’ active participation in physiotherapy depending on their (1) mental health problems, (2) physical difficulties, (3) level of cognitive function, (4) personality, (5) activities and participation, (6) age, (7) human environment, and (8) type of rehabilitation service where the individual undergoes treatment. For example, in cases where an individual lost self-confidence, participants offered practice tasks that the individual could achieve with little effort to make them experience success. Conversely, for individuals with overconfidence, participants would provide them with a relatively difficult practice task to help them realize the necessity of practice through the experience of failure. The interviews also revealed (9) motivational strategies used regardless of the individual’s condition. For instance, patient-centred communication was used to build a rapport with individuals, irrespective of their condition.

**Conclusion:** This qualitative study is the first to demonstrate motivational strategies that physiotherapists use based on the individual’s condition. Our findings provide a deeper understanding of the selection of motivational strategies in stroke rehabilitation.

## INTRODUCTION

Physical activity and exercise help individuals with stroke improve motor and cognitive impairment, increase independence in activities of daily living, and reduce the risk of stroke recurrence.^1, 2^ However, physical inactivity after stroke is highly prevalent.^3, 4^ Lack of motivation is a barrier to physical activity and exercise training in individuals with stroke.^5, 6^ Therefore, physiotherapists, who are primary healthcare professionals have great potential for promoting physical activity,^7^ are required to have the expertise in enhancing motivation in individuals undergoing rehabilitation. The addition of motivational strategies to physiotherapy has also been reported to increase adherence to exercise, have a positive effect on long-term exercise behaviour, improve self-confidence, and reduce levels of activity limitation in individuals with a range of chronic diseases.^8^

Motivation for physical activity and exercise is influenced by various factors.^5, 6, 9^ For example, cognitive and physical impairments are factors that decrease motivation,^9-11^ while self-confidence and support from family are the perceived motivators to physical activity.^9, 12, 13^ We previously reported that rehabilitation professionals used a range of motivational strategies to stroke rehabilitation based on comprehensive consideration of the individuals’ health-related information, suggesting that they use individual-tailored motivational strategies.^14, 15^ However, to our knowledge, it remains to be established how physiotherapists select their motivational strategies for each individual. Rehabilitation professionals tend to acquire skills to motivate individuals through their clinical experience.^14^ Therefore, qualitative exploration of the perspectives and experiences of expert physiotherapists in motivating individuals with stroke may help to better facilitate their participation in rehabilitation.

Qualitative research has been employed to explore factors influencing motivation in individuals with stroke.^6, 9^ Qualitative methods highlight the opinions of individuals to gain a deeper understanding of their perspectives and experiences within personal contexts that quantitative studies cannot explore or address.^16^ Therefore, qualitative methods may be highly beneficial for understanding how physiotherapists tailor motivational strategies toward each individual with stroke.

Therefore, the research question for this qualitative study was:

How do physiotherapists use different motivational strategies for individuals in stroke rehabilitation?

## METHODS

### Design

In our qualitative study, we used semi-structured interviews, which allow the interviewer and interviewee to diverge in order to pursue an idea or response in greater detail.^16^ We followed the Consolidated Criteria for Reporting Qualitative Research (COREQ) guidelines.^17^

### Participants, recruitment, and sampling strategies

A criterion sampling technique was used to select the participants.^18, 19^ To obtain rich and thick data, researchers need to recruit participants who are knowledgeable and can articulate and reflect on the research topic.^18^ Thus, the participants in this study were expected to be physiotherapists with expertise in stroke rehabilitation and with great interest in motivation strategies. To recruit participants, we first selected 66 potential physiotherapists from those who participated in our previous studies on motivational strategies for stroke rehabilitation^14, 15^ and had at least 10 years of clinical experience. E-mail invitations were sent to three or four potential participants every week until the required number of participants was reached. The potential participants to whom we sent the invitations were determined by computer-generated random numbers. Once interest was confirmed, participants were provided with an information sheet detailing the purpose of the study and information regarding data confidentiality alongside an informed consent form to be completed prior to interview conduction. Participants were informed that they could withdraw from the study at any time. They were also reimbursed for their time with a 5,000-yen gift card (approximately U.S. $45.00). The reimbursement for the participants was cited in the project description submitted to the ethical committee.

The final sample size was determined by the principle of data saturation.^20, 21^ Data saturation was considered as the point at which similar responses were elicited in the interviews with repeating rather than novel ideas. Qualitative studies require a minimum sample size of at least 12 participants to reach data saturation.^21^ Assuming that the data from approximately 20% of the participants would be excluded from analysis, we aimed to recruit a total of 15 participants.

### Data collection

The interviews were conducted from October 2020 to January 2021. An interview guide was developed based on our previous studies.^14, 15^ Prior to data collection, we carried out a pilot test to confirm that the interview guide was clear, understandable and capable of answering the research questions.^16^ The pilot test was conducted with three physiotherapists who had less than 10 years of clinical experience, resulting in minor changes to wording.

The term “motivation” in the present study was defined as “mental functions that produce the incentive to act; the conscious or unconscious driving force for action,” which was taken from the International Classification of Functioning, Disability and Health (ICF).^22^ Motivational strategy was defined as concrete tactics, techniques, or approaches to orient rehabilitation of individuals.^15^ These definitions were explained to participants prior to the interview to provide a common understanding of the terms. The interviewer orally asked the participants to answer questions based on their clinical experience and explained to them that there were no right or wrong answers to the interview questions. The interviews began with general questions about the most impressive experience of motivating an individual during rehabilitation followed by specific questions about which motivational strategies the participants used based on the individual’s condition (Table 1). In the specific questions, we comprehensively investigated the types of conditions of the individual that the participants thought needed to be motivated based on the ICF model: health condition, body functions and structures, activities, and participation, personal factors, and environmental factors.

**Table 1.** Semi-structured interview guide

| Topic | Question |
| --- | --- |
| General questions | ● Could you tell me your most impressive experience of motivating your individuals during stroke rehabilitation?<br>● Why did you choose that motivational strategy? |
| Motivational strategies to be used in consideration of health conditions of individuals | ● Regarding health condition domain of the ICF, what are the individual's conditions that you consider in deciding how to motivate individuals?<br>● Could you tell me how you have motivated the individuals with the conditions that you mentioned? |
| Motivational strategies to be used in consideration of body functions and structures of individuals | ● Regarding body functions and structures domain of the ICF, what are the individual's conditions that you consider in deciding how to motivate individuals?<br>● Could you tell me how you have motivated individuals with the condition that you mentioned? |
| Motivational strategies to be used in consideration of activities and participation of individuals | ● Regarding activities and participation domains of the ICF, what are the individual's conditions that you consider in deciding how to motivate individuals?<br>● Could you tell me how you have motivated individuals with the condition that you mentioned? |
| Motivational strategies to be used in consideration of personal factors of individuals | ● Regarding personal factors domain of the ICF, what are the individual's conditions that you consider in deciding how to motivate individuals?<br>● Could you tell me how you have motivated individuals with the condition that you mentioned? |
| Motivational strategies to be used in consideration of the environmental factors of individuals | ● Regarding personal factors domain of the ICF, what are the individual's conditions that you consider in deciding how to motivate individuals?<br>● Could you tell me how you have motivated individuals with the condition that you mentioned? |

ICF, International Classification of Functioning, Disability, and Health

Each interview was conducted by one of two interviewers, KO and KS, who were physiotherapists with over ten years of clinical experience, and video-conferencing software (Zoom, Zoom Video Communications Inc., CA, USA) was used due to COVID-19 restrictions. Video-conferencing interviews were the closest alternative to face-to-face interviews and allowed participants to join the interview from a distance. To ensure privacy, interviewers and interviewees participated in the interview in a private room. Considering the psychological burden of the participants, interviewers told participants that the interview could be stopped immediately when any psychological burden arose during the interview. Interviews lasted from 90 to 120 minutes. During and after each interview, the interviewer ensured that the interviewee was not distressed by the interview.

### Data analysis

All interviews were digitally recorded using the recording function of the video-conferencing software and were transcribed verbatim by an external provider of transcription services (Tokyo Hanyaku Co., Ltd.). The collected data was analysed with thematic analysis involving six stages: becoming familiar with the data, generating initial codes, searching for themes, reviewing themes, defining, and naming themes, and producing the report.^23^ The first author generated the initial codes. Furthermore, the first and second authors made subthemes and themes through repeated discussion until a consensus was reached. The third and fourth authors served as methodological auditors to examine the credibility of the conceptual interpretation of the original data.^24^ To protect the identities of the participants, we used pseudonyms. In quotations, we used an ellipsis mark to indicate the omitted words. Square brackets were used to supply words omitted by the speaker. A qualitative data management software (MAXQDA 2020, VERBI Software, Berlin, Germany) was used to analyse and organize the data.

## RESULTS

### Flow of participants through the study

E-mail invitations were sent to a total of 48 potential participants. However, there was no response from 33 of them, consequently 15 participated in the study. The characteristics of the participants are shown in Table 2 and are described in more detail in Supplementary Table. Thirteen participants were male, and the mean years of clinical experience was 14.9 years (standard deviation 4.0). All participants worked at different institutions in Japan. Thirteen participants had worked in a convalescent rehabilitation for individuals with subacute stroke, eight of whom were working there at the time of the interviews (Supplementary Table). Eight participants had a personal relationship with their respective interviewer.

**Table 2.** Characteristics of participants

| Variable | Value |
| --- | --- |
| Sex, male/female | 12 (80)/3 (20) |
| Years of clinical experience, years | 14.9 ± 4.0 |
| Current primary practice setting |  |
| Phase of stroke recovery of the individuals, acute/subacute/chronic | 5 (33)/9 (60)/3 (20) |
| Rehabilitation service, in-hospital/convalescent/homecare/out-of-hospital | 5 (33)/8 (53)/2 (13)/0 (0) |
| Other practice experience |  |
| Phase of stroke recovery of the individuals, acute/subacute/chronic | 4 (27)/5 (33)/9 (60) |
| Rehabilitation service, in-hospital /convalescent/homecare/ out-of-hospital | 5 (33)/5 (33)/4 (27)/4 (27) |

Values are presented as number (%) and mean ± standard deviation.

### Themes

Nine themes emerged from the data: they are summarized with representative quotes in Table 3 and described below.

**Table 3.**
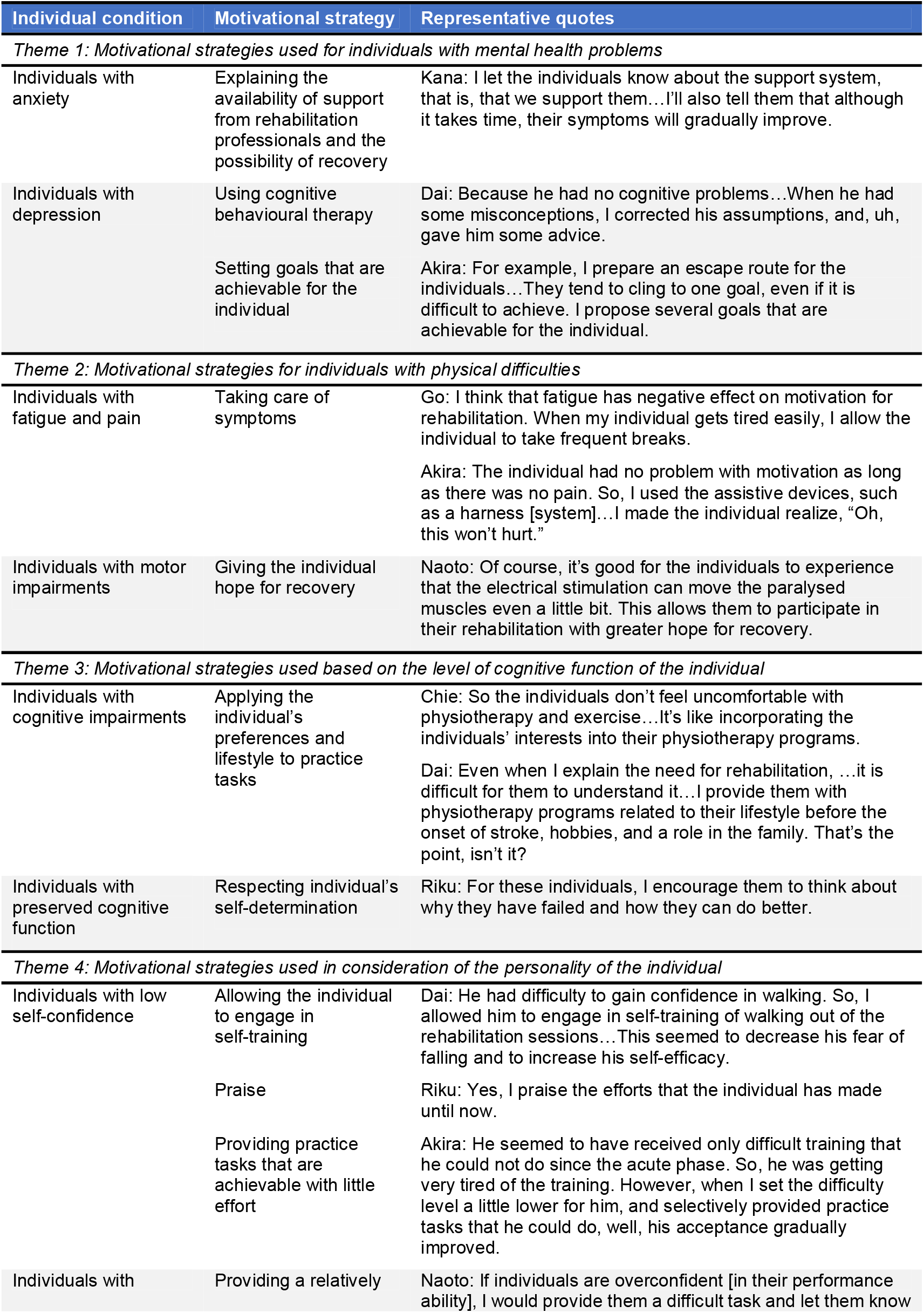
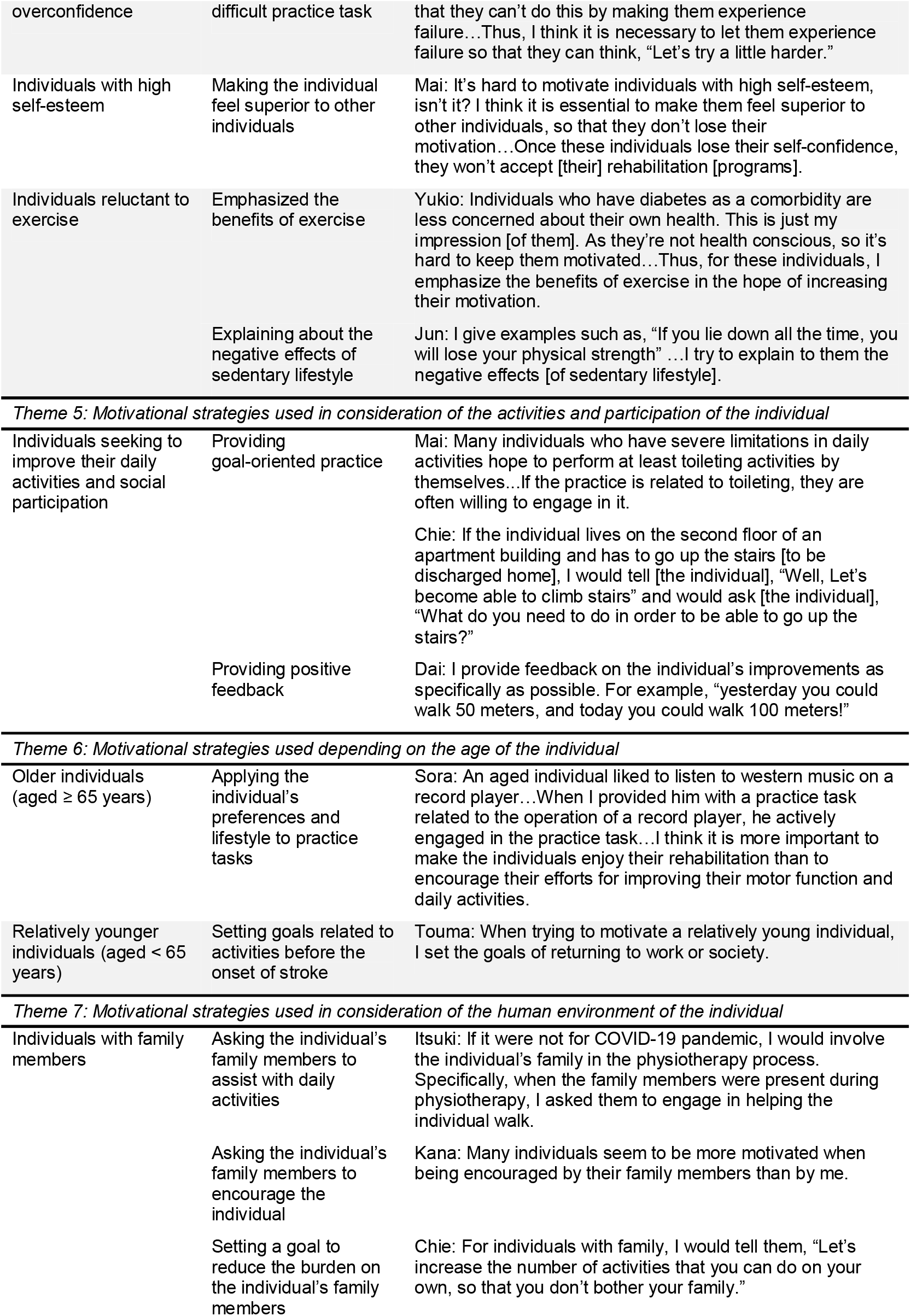
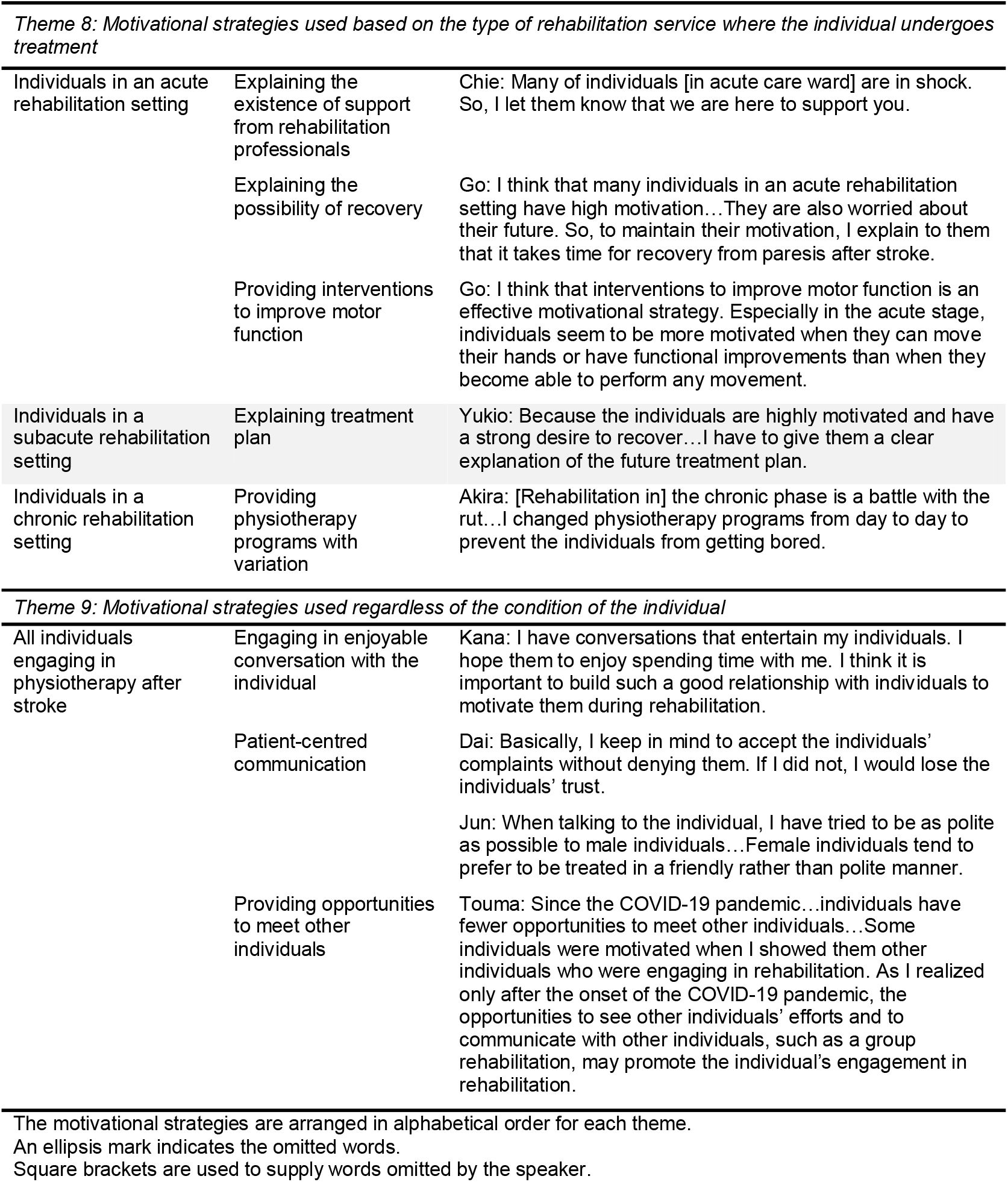
Motivational strategies used based on the condition of the individuals

#### Theme 1: Motivational strategies used for individuals with mental health problems

Participants thought that mental health problems, such as anxiety and depression, affected an individual’s motivation for physiotherapy. The participant Dai used cognitive behavioural therapy to motivate individuals with depression. He gave such individuals advice to correct their assumptions and misconceptions. The participant Akira also tried to increase their motivation by setting goals that were achievable for them:

> *For example, I prepare an escape route for the individuals…They tend to cling to one goal, even if it is difficult to achieve. I propose several goals that are achievable for the individual*.

In addition, if individuals were concerned about their future, the participant Kana provided them information on the availability of support from rehabilitation professionals and the possibility of recovery to reduce their anxiety:

> *I let the individuals know about the support system, that is, that we support them…I’ll also tell them that although it takes time, their symptoms will gradually improve*.

Thus, participants tried to encourage the active participation of the individuals in the rehabilitation program by reducing their depressive symptoms and anxiety.

#### Theme 2: Motivational strategies for individuals with physical difficulties

The analysis revealed that participants used different motivational strategies for individuals with fatigue, pain, and motor impairment. In cases where individuals presented fatigue and pain, the participants tried to maintain their motivation by addressing these symptoms, such as by taking frequent breaks and recommending the use of assistive devices. Additionally, for individuals with motor impairment, some participants tried to give individuals hope for recovery by using therapeutic electrical stimulation. The participant Naoto stated,

> *Of course, it’s good for the individuals to experience that the electrical stimulation can move the paralysed muscles even a little bit. This allows them to participate in their rehabilitation with greater hope for recovery*.

#### Theme 3: Motivational strategies used based on the level of cognitive function of the individual

Participants thought that when individuals could not understand the importance of practice and exercise due to cognitive impairment, it would be difficult to encourage their participation in physiotherapy programs. Thus, participants tried to prevent the individuals from being reluctant to participate in their physiotherapy by providing rehabilitation programs related to the individuals’ preferences and their lifestyle before the onset of stroke. In contrast, for individuals with preserved cognitive function, the participant Riku respected their self-determination and provided them with the opportunity to determine for themselves how to solve their problems.

#### Theme 4: Motivational strategies used in consideration of the personality of the individual

Participants used different motivational strategies depending on the degree of self-confidence of the individual. For individuals with low self-confidence, practice tasks achievable with little effort were offered to allow them to experience success. Additionally, allowing individuals to engage in self-training and praise were also considered effective in increasing self-confidence. In contrast, some participants mentioned that individuals with overconfidence needed to experience of failure to understand the necessary of practice. Thus, they provided these individuals with a relatively difficult practice task. Higher educated individuals and those with professional background such as a president of a company or a university professor were thought to tend to have higher self-esteem. To motivate such individuals, the participant Mai emphasized the importance of making them feel superior to other individuals:

> *It’s hard to motivate individuals with high self-esteem, isn’t it? I think it is essential to make them feel superior to other individuals, so that they don’t lose their motivation…Once these individuals lose their self-confidence, they won’t accept [their] rehabilitation [programs]*.

In motivating individuals who are reluctant or refuse to participate in physiotherapy programs, participants tried to make them understand the necessity of physiotherapy. For example, participants thought that individuals with comorbid diabetes mellitus tended to be less willing to engage in exercise therapy, because many of them were not health conscious and did not like to exercise by nature. To motivate individuals who were reluctant to exercise, participants emphasized the benefits of exercise to them. In addition, Jun explained the negative effects of a sedentary lifestyle:

> *I give examples such as, “If you lie down all the time, you will lose your physical strength”…I try to explain to them the negative effects [of sedentary lifestyle]*.

#### Theme 5: Motivational strategies used in consideration of the activities and participation of the individual

For individuals seeking to improve their daily activities and social participation, participants provided goal-oriented practice tasks to enhance their adherence to physiotherapy programs. For example, Mai reported,

> *Many individuals who have severe limitations in daily activities hope to perform at least toileting activities by themselves*…*If the practice is related to toileting, they are often willing to engage in it*.

Additionally, if individuals will be discharged to their home, participants provided the individuals practice tasks taking into consideration the individual home environment of the individual. Furthermore, positive feedback was used to help individuals be aware of their improvements.

#### Theme 6: Motivational strategies used depending on the age of the individual

Compared to relatively younger individuals (aged < 65 years), older individuals (aged ≥ 65 years) were thought to have less adherence to rehabilitation programs because of giving up on recovery and age-related physical weaknesses. Participants prioritized making their physiotherapy enjoyable rather than to encourage their efforts. In contrast, for relatively younger individuals, participants believed that providing practice tasks to help return to work could increase individual adherence to physiotherapy programs.

#### Theme 7: Motivational strategies used in consideration of the human environment of the individual

For individuals with family members, support from their family, such as encouragement and assistance with daily activities, was considered as an effective motivator. Some participants stated that setting a goal of reducing the burden on family members could encourage their participation in physiotherapy programs.

#### Theme 8: Motivational strategies used based on the type of rehabilitation service where the individual undergoes treatment

Participants varied their motivational strategies depending on the different rehabilitation services where individuals underwent treatment, such as acute, subacute, and chronic rehabilitation settings. Some participants thought that individuals in an acute stroke rehabilitation setting were depressed or anxious. As mentioned above as a motivational strategy for individuals with anxiety, participants explained the availability of support from rehabilitation professionals and the possibility of functional recovery to these individuals. In addition, the participant Go stated that especially for individuals with acute stroke, interventions to improve motor function were an effective motivational strategy. The participant Yukio mentioned that individuals with subacute stroke, who were admitted to a convalescent rehabilitation hospital, were more motivated than those in acute or chronic rehabilitation settings. He emphasized the importance of explaining his treatment plan to maintain their proactiveness. Furthermore, participants felt that physiotherapy programs in a chronic rehabilitation setting tended to activities becoming habitual and uninteresting. Akira tried to prevent the individuals from getting bored by providing variations of physiotherapy programs.

#### Theme 9: Motivational strategies used regardless of the condition of the individual

The interviews revealed some motivational strategies that were commonly used regardless of the individual’s condition. Participants mentioned the importance of building a rapport with individuals to motivate them during physiotherapy. Patient-centred communication, which included eliciting the individual’s agenda with open-ended questions, not interrupting the individual, and engaging in focused active listening,^25^ seemed to be used as a strategy to establish a relationship of trust with the individuals. Engaging in enjoyable conversation with the individuals was also thought to help build a rapport with them.

Some participants thought that the opportunities to interact with other individuals may promote the individual’s engagement in physiotherapy. Touma stated,

> *Since the COVID-19 pandemic…individuals have fewer opportunities to meet other individuals…Some individuals were motivated when I showed them other individuals who were engaging in rehabilitation. As I realized only after the onset of the COVID-19 pandemic, the opportunities to see other individuals’ efforts and to communicate with other individuals, such as a group rehabilitation, may promote the individual’s engagement in rehabilitation*.

## DISCUSSION

To the best of our knowledge, this qualitative research is the first to describe the motivational strategies used for physiotherapy according to the condition of the individuals after a stroke. Consistent with our previous studies,^14, 15^ participants decided which motivational strategies to use based on comprehensive consideration of the individual’s health-related information including their health condition, body functions and structures, activities, and participation, personal factors, and environmental factors. Our results suggest that physiotherapists use different motivational strategies for each individual based on their mental health conditions, physical difficulties, the level of cognitive function, personality, activities and participation, age, human environment, and the type of rehabilitation service where the individual undergoes treatment. The qualitative findings provide a deeper understanding of the selection of motivational strategies in stroke rehabilitation.

The participants selected the motivational strategy to use in consideration of the cause of an individual’s lack of motivation. For example, anxiety hampered the stroke rehabilitation effort and prevented individuals from returning to their usual activities.^26^ Participants tried to reduce individual anxiety for their future by providing information on the availability of support programs from rehabilitation professionals and the possibility of recovery. Information provision by involving individual or providing reinforcement may reduce anxiety in individuals with stroke, although the effect size is small.^27^ Participants also tried to motivate their individuals by respecting each individual’s values, preferences, experiences, and self-determination. These strategies tailored to the individual to promote and support patient-centred care. Patient-centred care has been shown to promote positive effects on physical activity, occupational performance and satisfaction, autonomy, and rehabilitation satisfaction in individuals with stroke.^28^ Thus, the use of different motivational strategies according to the individual’s condition appears to not only enhance adherence, but also to improve rehabilitation outcomes.

The interviews also helped to identify motivational strategies commonly applied to all individuals undergoing physiotherapy after stroke, regardless of the condition of the individual. The interviews suggested that a rapport with therapists and individuals was essential to enhance an individual’s active participation in physiotherapy. Establishing a collaborative partnership between the therapist and the individual is also a patient-centred intervention.^28^ Danzl et al.^29^ reported four strategies to build rapport and trust as follows: 1) spending time with the individuals without rushing them, 2) developing a deeper understanding of the individuals, 3) using a sense of humour, when appropriate, and 4) showing compassion, empathy, and respect. Listening to the individual while refraining from interrupting, understanding the individual’s perspective, and expressing empathy are key features of patient-centred communication.^25^ A sense of humour contributed to making the individual enjoy the rehabilitation. Thus, patient-centred communication and engaging in enjoyable conversation used by the participants in this study may have been used to effectively establish a relationship of trust with the individual. Moreover, we found that providing opportunities for individuals to interact with each other is also effective in motivating individuals for rehabilitation, irrespectively of their condition. Interacting with other individuals, such as watching other individuals’ efforts and communicating with other individuals, has been shown to be a perceived motivator to physical activity in individuals with stroke.^6, 9^ However, as the participant Touma mentioned, the opportunities for individuals to interact with each other have diminished since the onset of the COVID-19 pandemic. If the COVID-19 pandemic continues, it will become increasingly important to provide an environment that allows individuals to safely interact with each other.

As a note of caution, most of the participants had experience working in a convalescent rehabilitation centre for individuals with subacute stroke. Thus, the perspectives and experiences of motivating individuals in a subacute rehabilitation setting might be overstated in the data. To minimize this sampling bias, we recommended that further studies stratify the participants by the individual’s phase of stroke recovery in which they most frequently worked and the rehabilitation services where they worked, such as in-hospital, out-of-hospital, and homecare rehabilitation.

In conclusion, this qualitative study suggests that physiotherapists use different strategies depending on the individual’s mental health conditions, physical problems, the level of cognitive function, personality, activities and participation, age, human environment, and the type of rehabilitation service where the individual undergoes treatment to motivate individuals with stroke during physiotherapy. These findings can provide an experience-based recommendation regarding the selection of motivational strategies in stroke rehabilitation.

## Data Availability

All data produced in the present study are available upon reasonable request to the corresponding author.

## Acknowledgements

We thank Anna, from Editage Group (www.editage.jp), for editing a draft of this manuscript.

## Supplementary Material

**Supplementary Table.** Participants’ characteristics

| Pseudonym | Sex | Years of clinical experience | Current primary practice setting | Other practice experience |
| --- | --- | --- | --- | --- |
| Akira | Male | 13 | Convalescent rehabilitation for individuals with subacute stroke | In-hospital rehabilitation for individuals with acute stroke<br>Out-of-hospital rehabilitation for individuals with chronic stroke |
| Chie | Female | 12 | Convalescent rehabilitation for individuals with subacute stroke | Homecare rehabilitation for individuals with chronic stroke |
| Dai | Male | 16 | Convalescent rehabilitation for individuals with subacute stroke | In-hospital rehabilitation for individuals with acute stroke |
| Eito | Male | 11 | Convalescent rehabilitation for individuals with subacute stroke | Not applicable |
| Go | Male | 15 | In-hospital rehabilitation for individuals with acute stroke | In-hospital rehabilitation for individuals with chronic stroke |
| Haruto | Male | 11 | In-hospital rehabilitation for individuals with acute stroke | Convalescent rehabilitation for individuals with subacute stroke<br>Homecare rehabilitation for individuals with chronic stroke |
| Itsuki | Male | 14 | Convalescent rehabilitation for individuals with subacute stroke | Out-of-hospital rehabilitation for individuals with chronic stroke |
| Jun | Male | 11 | Homecare rehabilitation for individuals with chronic stroke | Convalescent rehabilitation for individuals with subacute stroke<br>In-hospital rehabilitation for individuals with acute stroke |
| Kana | Female | 25 | In-hospital rehabilitation for individuals with acute, subacute, and chronic stroke | Not applicable |
| Mai | Female | 20 | Convalescent rehabilitation for individuals with subacute stroke | Homecare rehabilitation for individuals with chronic stroke |
| Naoto | Male | 20 | Convalescent rehabilitation for individuals with subacute stroke | Not applicable |
| Riku | Male | 13 | In-hospital rehabilitation for individuals with acute stroke | Convalescent rehabilitation for individuals with subacute stroke<br>Out-of-hospital rehabilitation for individuals with chronic stroke |
| Sora | Male | 13 | Convalescent rehabilitation for individuals with subacute stroke | Out-of-hospital rehabilitation for individuals with chronic stroke |
| Touma | Male | 15 | In-hospital rehabilitation for individuals with acute stroke | Convalescent rehabilitation for individuals with subacute stroke<br>Homecare rehabilitation for individuals with chronic stroke |
| Yukio | Male | 15 | Homecare rehabilitation for individuals with chronic stroke | Convalescent rehabilitation for individuals with subacute stroke<br>In-hospital rehabilitation for individuals with acute stroke |

The pseudonyms of participants are arranged in alphabetical order.

